# Role of favipiravir in the treatment of adult patients with moderate to severe COVID-19: a single-center, prospective, observational, sequential cohort study from Hungary

**DOI:** 10.1101/2020.11.26.20238014

**Authors:** Balint Gergely Szabo, Katalin Szidonia Lenart, Borisz Petrik, Zsofia Gaspar, Zsofia Balogh, Zsuzsanna Banyai, Emese Banyasz, Jozsef Budai, Eszter Czel, Katalin Fried, Adrienn Hanuska, Noemi Kiss-Dala, Csaba Lorinczi, Krisztina Nemesi, Janos Kadar, Eva Livia Nagy, Akos Osvald, Edina Petrovicz, Alexandra Riczu, Judit Szanka, Beata Szathmary, Andrea Szombati, Szilvia Toth, Zsuzsanna Varnai, Orsolya Woller, Janos Szlavik, Botond Lakatos

## Abstract

**Background:** Preliminary data suggests that favipiravir might have a role in COVID-19 treatment. Our aim was to assess the role of favipiravir in the treatment of COVID-19.

**Methods:** A single-center, prospective, observational, sequential cohort study was performed among consecutive adults hospitalized with PCR-confirmed COVID-19 between March– July,2020. Patients were screened for inclusion by *a priori* criteria, and were included in the favipiravir cohort if SOC+FVP, or the non-favipiravir group if SOC±other antiviral medications without FVP were administered for >48 hours. Treatment allocation was done per national guidelines. For COVID-19 diagnosis and severity, ECDC and WHO definitions were utilized, and daily *per protocol* hospital follow-up was done. Primary composite end-point was disease progression (14-day all-cause death, need for mechanical ventilation, or immunomodulatory therapy). For statistical comparison, Fisher’s exact test and Mann– Whitney U-test were used.

**Results:** In all, 75 patients were included per cohort. In the FVP cohort, chronic heart disease (36/75, 48.0% vs. 16/75, 21.3%, p<0.01) and diabetes mellitus (23/75, 30.7% vs. 10/75, 13.3%, p<0.01) were more prevalent, hospital LOS (18.5±15.5 days vs. 13.0±8.5 days, p<0.01) was higher. Disease progression (17/75, 22.7% vs. 10/75, 13.3%, p=0.13), 14-day all-cause death (9/75, 12.0% vs. 10/75, 13.3%, p=0.8) and need for mechanical ventillation (8/75, 10.7% vs. 4/75, 5.3%, p=0.22) were similar between groups. Immunomodulatory therapies were administered frequently among patients receiving FVP (10/75, 13.3% vs. 1/75, 1.3%, p<0.01).

**Conclusions:** In this study, favipiravir did not seem to affect disease progression. Further data are needed to position this drug among the anti-SARS-CoV-2 armamentarium.

## 1. INTRODUCTION

As the pandemic caused by severe acute respiratory syndrome coronavirus-2 (SARS-CoV-2) is ongoing, investigators are searching for therapeutic strategies to cure coronavirus disease-19 (COVID-19). As of September 2020, the number antiviral drugs proven to effectively inhibit viral replication is low [1]. Furthermore, it is not known which antivirals are unequivocally able to attenuate the pathological progression of COVID-19 in most patients with different severity. Favipiravir (FVP) is an antiviral pyrazinecarboxamide derivative, licensed for influenza in Japan. Data from the literature suggested that favipiravir might also be useful in COVID-19 treatment [2, 3]. Therefore, our goal was to assess the effect of favipiravir in the treatment of adult patients hospitalized at our centre with moderate to severe COVID-19.

## 2. METHODS

### 2.1 Study design and settings

A single-center, prospective, observational, sequential cohort study was performed among adults diagnosed and hospitalized with COVID-19 at our centre between March 15 – July 15, 2020 (first COVID-19 case was confirmed on March 4 in Hungary). Our centre is a national-level referreal institution, and the main COVID-19 centers of the country with >150 dedicated beds. The study was in accordance with the Helsinki Declaration, as well as national ethical standards. The institutional review board approved the study protocol, and informed consent was obtained from every patient.

### 2.2 Patient eligibility and inclusion

Consecutive adult (≥18 years of age) patients hospitalized at our centre during the study period with respiratory SARS-CoV-2 polymerase chain reaction (PCR) confirmed COVID-19 cases, irrespective of COVID-19 disease severity, were eligible for inclusion. To overcome selectional bias, all patients were prospectively screened for inclusion by our COVID-19 team (composed of attending physicians), by daily real-time visits at the ward and intensive care unit (ICU), and case assessment using hospital electronic records. Selection was done by using *a priori* inclusion/exclusion criteria. Inclusion criteria were: 1) PCR based confirmation of COVID-19 diagnosis in a clinically compatible case, with any disease severity; 2) received standard of care (SOC) or any of the exposure treatments for >48 hours after diagnosis. Exclusion criteria were: 1) the patient was intubated, died or discharged within ≤48 hours after diagnosis; 2) received SOC or any of the exposure treatments for ≤48 hours after diagnosis; 3) received any antiviral medication before diagnosis 4) patient data was not accessible through hospital electronic database.

Included patients were grouped into two sequential cohorts in 1:1 fashion, according to treatment exposures: favipiravir (FVP) cohort consisted of patients receiving SOC+FVP, non-favipiravir (non-FVP) cohort included patients who were administered SOC±other antiviral medications. Other possible antiviral medications were chloroquine, hydroxychloroquine, lopinavir/ritonavir, remdesivir and oseltamivir (please refer to the adequate section for details).

### 2.3 Data collection

A dedicated database was established for the study aim by manual extraction of data from hospital electronic records and written charts. Data was transferred anonymously to an *a priori* standardized case report form. Data collected: 1) age and gender; 2) comorbidities; 3) intensive care unit (ICU) admission during hospitalization; 4) length of stay (LOS) and ICU LOS; 4) clinical parameters at COVID-19 diagnosis (symptom onset, COVID-19 severity, supplementary oxygen demand, peripheral oxygen saturation, acute respiratory distress syndrome [ARDS], cytokine storm [CS], acute respiratory failure); 5) laboratory parameters at diagnosis (blood leucocyte, absolute neutrophil granulocyte, lymphocyte and platelet counts, hemoglobin, CRP, procalcitonin, serum ferritin, high sensitivity cardiac Troponin-I, serum interleukin-6, serum creatinine, LDH and D-dimer); 6) microbiological and radiological parameters at diagnosis (blood cultures and chest computed tomography [CT]); 7) characteristics of antimicrobial therapies and supportive care; 8) characteristics of immunomodulatory therapies; 9) study outcomes. Variables with ≥5% of missing measurements were omitted from final analysis, no imputation was seeked.

### 2.4 Diagnostic and therapeutic strategies at our centre during the pandemic

At our centre, we follow the *European Centre for Disease Prevention and Control* (ECDC) COVID-19 case definition for diagnosis ascertainment [4]. A clinically suspicious case presentation (cardinal symptoms: fever, dyspnea, cough) is confirmed if a respiratory specimen is positive for SARS-CoV-2 nucleic acid by PCR. Respiratory specimens are taken by a team of trained nurses with oro-/nasopharyngeal sampling in spontaneously breathing patients, or by mini-BAL in ventilated patients. Disease severity is determined by *World Health Organization* (WHO) criteria [5]. Disease onset is the first day of symptom apperance as reported by the patient, or day of first PCR positivity if symptom appearance could not be reported. The day of first SARS-CoV-2 PCR positivity is given as COVID-19 diagnosis day. Acute respiratory failure and ARDS were defined following the 2012 Berlin criteria, CS is diagnosed by trends of clinical and laboratory parameters and HScore, as proposed by *Mehta et al*. [6, 7].

At our centre, COVID-19 patient care is facilitated by standardized in-house diagnostic and therapeutic strategies, composed of written protocols and check-lists updated since March 2020 according to literature evidence. Physical examination, laboratory studies, arterial blood gas analyses are done daily. Imaging studies (chest X-ray and/or chest CT) are done on the day of COVID-19 diagnosis, and days +7, +14, and if disease progression (persistent fevers for ≥72 hours, deteriorating hypoxaemia) or clinical instability (newly onset dyspnea, chest pain, hypotension, altered mentation) is documented. Every febrile (tympanal temperature ≥37.8°C) patient has 2 sets of blood cultures taken. Testing for SARS-CoV-2 PCR negativity is initiated 24 hours after defervescence and clinical stability. Respiratory samples for SARS-CoV-2 PCR are collected every 48 hours, PCR negativity is documented on the day when two consecutive samples are proven to be negative. Daily follow-up of patients is done until death or hospital discharge, post-discharge follow-up is not routinely arranged. Microbiological diagnostics are performed at the Microbiology Laboratory of our centre.

COVID-19 antiviral therapies are allocated *per protocol*, in an open-label and non-randomized fashion (excluding patients in randomized clinical trials), but allocation is affected by drug availability on a national level. At our centre, treatment allocation is done in accordance with the “Hungarian Coronavirus Handbook” [8]. Before favipiravir introduction to Hungary (May 15, 2020), all patients received other antiviral medications: chloroquine (1g loading dose, 1×500mg maintenance thereafter, minimum of 7 days) or hydroxychloroquine (2×400mg loading dose, 2×200mg maintenance thereafter, min. of 5 days) or lopinavir/ritonavir (200/50mg in 2×2 capsules, min. of 7 days), depending on patient contraindications and drug availability. As the influenza season was overlapping with the first month of the epidemic in Hungary, oseltamivir (2×75mg) was also administered to patients, pending influenza A/B respiratory PCR results. Remdesivir was not available in Hungary during the study period. After favipiravir distribution, all patients received it *per protocol* in monotherapy at our centre (2×1600mg loading dose, 2×600mg maintenance thereafter, min. of 10 days). Patients are routinely receiving SOC (on-demand oxygen therapy, respiratory support, intravenous fluid replenishment, antipyretics, antitussive and bronchodilator drugs). Empirical antibiotics per *Infectious Diseases Society of America* (IDSA) community-acquired pneumonia guideline are administered if disease progression or clinical instability is documented, immunomodulatory agents are administered in a clinical study to patients with CS or critical COVID-19, per COVID-19 team initiation [9].

### 2.5 Outcomes

The primary endpoint was disease progression, a negative composite of any of the following: 1) 14-day all-cause death, 2) need for mechanical ventilation, 3) need for immunomodulatory therapy for COVID-19 at ward or ICU. 14-day all-cause death was defined as exitus within 14 days from COVID-19 diagnosis during hospital stay. Need for mechanical ventilation was defined as a completed endotracheal intubation in relation to COVID-19, per decision of an ICU crash team. Need for immunomodulatory therapy was defined if any immunomodulatory drug was initiated at any dose, suggested by the “Hungarian Coronavirus Handbook” (at our centre, depending on availability: tocilizumab, ruxolitinib, baricitinib, intravenous immunoglobulin, reconvalescent plasma or systemic corticosteroids), excluding systemic corticosteroids started for alternative causes.

Secondary endpoints were 14-day all-cause mortality, need for mechanical ventilation, need for immunomodulatory therapy, rate of PCR negativity (at the end of hospitalization or sooner). Time analyses were done between cohorts by comparing time intervals from diagnosis day to disease progression, to all-cause mortality, to mechanical ventilation, to immunomodulatory therapy, to PCR negativity.

### 2.6 Statistical analysis

Continuous variables are expressed as median±interquartile range (IQR). Normality was checked with the *Shapiro–Wilks*-test. Categorical variables are expressed as absolute numbers (n) with relative percentages (%). Statistical comparisons were done with *Mann– Whitney* U-test *or Fisher*’s exact test. For identification of independent risk factors associating with disease progression, uni- and multivariate binomial logistic regression was performed. All plausible parameters and those with a *p*-value of ≤0.1 in univariate analysis were entered into a forward stepwise multivariate logistic regression (entry criterion: *p*=0.05, removal criterion: *p*=0.1). Maximal number of predictors was estimated with the common 1:10 rule-of-thumb, goodness-of-fit was tested by the *Hosmer–Lemeshow* test. For all tests, a two-tailed *p*-value of < 0.05 determined statistical significance. Data collection was completed with Microsoft Office Excel 2016, tests were calculated using IBM SPSS Statistics 23. For reporting, we adhere to the *Strengthening the Reporting of Observational studies in Epidemiology* (STROBE) Statement [10].

## 3. RESULTS

### 3.1 Baseline and clinical characteristics

In all, 150 patients were enrolled during the study period, 75 in both cohorts. Baseline and clinical characteristics are described in *Table 1*. Median age was 66.0±12.4 years, with representation of older patients in the FVP group (71.5±15.1 vs. 61.0±21.5 years, p<0.01). Genders and most comorbidities were equally distributed between cohorts, essential hypertension (57/75, 76.0% vs. 38/75, 50.7%, p<0.01) chronic heart disease (36/75, 48.0% vs. 16/75, 21.3%, p<0.01), diabetes mellitus (18/75, 24.0% vs. 6/75, 8.0%, p=0.01) and chronic cerebral disease (23/75, 30.7% vs. 10/75, 13.3%, p<0.01) were more prevalent in the FVP cohort. At diagnosis, 35/75 (53.3%) and 41/75 (54.7%) patients had severe disease in the FVP and non-FVP cohorts, respectively (p=0.41), and ARDS or CS was not documented. Need for oxygen supportation was needed in similar rates (27/75, 36.0% vs. 21/75, 28.5%, p=0.29). Documented bloodstream-infections were rare (1/75, 0.7%). Patients in the FVP group had higher white blood cell (6.5±4.2×10^9^/L vs. 5.5±3.3×10^9^/L, p=0.04), platelet counts (225±117×10^9^/L vs. 191±108×10^9^/L, p=0.03), serum creatinine (86.0±49.3 μmol/L vs. 71.0±37.5 μmol/L, p=0.03) and D-dimer (1163±2038 ng/mL vs. 850±479 ng/mL, p=0.03), lower hemoglobin levels (117±24 g/l vs. 131±20 g/L, p<0.01). The rate of chest CT positivity did not differ between cohorts (54 / 64, 84.4% vs. 13 / 14, 92.8%, p=0.67).

**Table 1.** Baseline and clinical characteristics of adult COVID-19 patients included in the study, grouped by favipiravir administration.

| PARAMETER | Total<br>(n=150) | FVP cohort<br>(n=75) | Non-FVP cohort<br>(n=75) | p value |
| --- | --- | --- | --- | --- |
| Age (years, median±IQR, min–max) | 66.0±12.4 (20.0–93.0) | 71.5±15.1 (24.0–93.0) | 61.0±21.5 (20.0–91.0) | <0.01 |
| Male gender (n, %) | 76 (50.7) | 37 (49.3) | 39 (52.0) | 0.74 |
| ICU admission during hospitalization (n, %) | 17 (11.3) | 12 (16.0) | 5 (6.7) | 0.07 |
| Time from disease onset to antiviral therapy<br>(days, median±IQR, min–max) | 3.0±6.0 (1–34) | 2.0±3.0 (1–34) | 7.5±9.0 (1–21) | <0.01 |
| Time from positive PCR to antiviral therapy<sup>a</sup><br>(days, median±IQR, min–max) | 1.0±2.0 (1–31) | 1.0±2.0 (1–31) | 1.0±2.0 (1–6) | 0.88 |
| Comorbidities (n, %): |  |  |  |  |
| - Essential hypertension | 95 (63.3) | 57 (76.0) | 38 (50.7) | <0.01 |
| - Chronic heart disease | 52 (34.7) | 36 (48.0) | 16 (21.3) | <0.01 |
| - Chronic vascular disease | 14 (9.3) | 10 (13.3) | 4 (5.3) | 0.1 |
| - Chronic pulmonary disease | 20 (13.3) | 11 (14.7) | 9 (12.0) | 0.6 |
| - Chronic renal disease | 9 (6.0) | 7 (9.3) | 2 (2.7) | 0.09 |
| - Chronic hepatic disease | 5 (3.3) | 4 (5.3) | 1 (1.3) | 0.18 |
| - Chronic cerebral disease | 24 (16.0) | 18 (24.0) | 6 (8.0) | 0.01 |
| - Diabetes mellitus | 33 (22.0) | 23 (30.7) | 10 (13.3) | <0.01 |
| - Active oncological malignancy | 21 (14.0) | 15 (20.0) | 6 (8.0) | 0.05 |
| - Active hematological malignancy | 13 (8.7) | 9 (12.0) | 4 (5.3) | 0.16 |
| - Systemic autoimmune disease | 7 (4.7) | 2 (2.7) | 5 (6.7) | 0.16 |
| - Chronic systemic corticosteroid treatment | 2 (1.3) | 2 (2.7) | 0 | 0.14 |
| - Chronic systemic immunosuppressive treatment | 2 (1.3) | 0 | 2 (2.7) | 0.23 |
| - Smoking | 2 (1.3) | 2 (2.7) | 0 | 0.16 |
| Clinical characteristics at COVID-19 diagnosis: |  |  |  |  |
| - Peripheral oxygen saturation (%; median±IQR, min–max) | 95±7 (51–99) | 94±7 (51–99) | 95±7 (78–99) | 0.94 |
| - Need for oxygen supportation (n, %) | 48 (32.0) | 27 (36.0) | 21 (28.0) | 0.29 |
| - ARDS (n, %) | 0 | 0 | 0 | n.a. |
| - Acute respiratory failure (n, %) | 1 (0.7) | 0 | 1 (1.3) | n.a. |
| - CS (n, %) | 0 | 0 | 0 | n.a. |
| Severity of COVID-19<sup>b</sup> (n, %): |  |  |  |  |
| - Non-severe disease | 74 (49.3) | 40 (53.3) | 34 (45.3) | 0.41 |
| - Severe disease | 76 (50.7) | 35 (46.7) | 41 (54.7) |  |
| Laboratory characteristics at COVID-19 diagnosis<br>(median±IQR, min–max): |  |  |  |  |
| - White blood cell count (x10<sup>9</sup>/L) | 6.2±3.7 (2.3–21.7) | 6.5±4.2 (2.7–21.7) | 5.5±3.3 (2.3–16.4) | 0.04 |
| - Neutrophil granulocyte count (x10<sup>9</sup>/L) | 4.0±2.9 (0.4–20.1) | 4.2±2.9 (0.4–20.1) | 3.8±3.6 (1.6–14.9) | 0.09 |
| - Lymphocyte count (x10<sup>9</sup>/L) | 1.2±0.7 (0.3–3.7) | 1.2±0.8 (0.3–3.7) | 1.1±0.7 (0.3–2.6) | 0.07 |
| - Platelet count (x10<sup>9</sup>/L) | 203±93 (12–743) | 225±117 (12–743) | 191±108 (78–414) | 0.03 |
| - Hemoglobin level (g/L) | 124±32 (67–163) | 117±24 (67–152) | 131±20 (89–163) | <0.01 |
| - C-reactive protein (mg/L) | 41±94 (1–316) | 37±90 (1–229) | 44.7±91.9 (1–316) | 0.52 |
| - Procalcitonin (µg/L) | 0.0±0.1 (0–92.8) | 0.1±0.1 (0–92.8) | 0.0±0.1 (0–14.4) | 0.08 |
| - Interleukin-6 (pg/mL) | 16.1±20.0 (2.7–76.9) | 13.9±22.3 (2.7–76.9) | 20.0±10.6 (15.6–46.0) | 1.0 |
| - Serum ferritin (µg/L) | 397±520 (37–6001) | 403±555 (37–6001) | 364±219 (84–1268) | 0.72 |
| - Cardiac Troponin-I (ng/mL) | 0.012±0.026 (0.001–2.171) | 0.012±0.025 (0.001–2.171) | 0.008±0.02 (0.002–0.056) | 0.37 |
| - Serum creatinine (µmol/L) | 75.0±35.5 (36.0–470.0) | 86.0±49.3 (36.0–446.0) | 71.0±37.5 (37.0–470.0) | 0.03 |
| - Serum LDH (IU/L) | 512±242 (214–2240) | 491±232 (214–2240) | 534±275 (241–1370) | 0.51 |
| - Serum D-dimer (ng/mL) | 1111±1405 (160–49937) | 1163±2038 (160–49937) | 850±479 (316–1319) | 0.03 |
| Blood cultures taken per patient (n, %) | 76 (50.7) | 36 (48.0) | 40 (53.3) | 0.46 |
| Detected bloodstream-infections (n, %) | 1 (0.7) | 1 (1.3) | 0 | 0.31 |
| Chest CT positive for COVID-19 / chest CT obtained | 67 / 78 (85.9) | 54 / 64 (84.4) | 13 / 14 (92.8) | 0.67 |

<sup>a</sup> From first positive SARS-CoV-2 PCR, sampled from the respiratory tract
<sup>b</sup> Per *World Health Organization* criteria
n.a. Not applicable.

### 3.2 Outcomes, therapeutic approaches

Outcomes and therapeutic approaches are detailed in *Table 2*. Disease progression showed no statistically significant difference (17/75, 22.7% vs. 10/75, 13.3%, p=0.13) between cohorts. Rates of 14-day all-cause mortality (9/75, 12.0% vs. 10/75, 13.3%, p=0.8) and need for mechanical ventilation (8/75, 10.7% vs. 4/75, 5.3%, p=0.22) were also similar, need for immunomodulatory therapies was higher among patients receiving FVP (10/75, 13.3% vs. 1/75, 1.3%, p<0.01). FVP expuosure was not retained as an independent covariate in multivarite logistic regression modelling disease progression (*Table 3*.). Although there was a statistically non-significant tendency for higher PCR negativity rate (55/75, 73.0% vs. 43/75, 57.3%, p=0.05), median time to PCR negativity was significantly longer (22.0±15.5 days vs. 13.0±12.0 days, p<0.01) in the FVP cohort. Median time from diagnosis to disease progression (8.0±9.0 days vs. 4.5±9.8, p=0.08) and time to exitus (16.0±14.0 days vs. 8.5±10.3 days, p=0.03) were both longer among patients receiving FVP.

**Table 2.** Outcomes and therapeutic approaches to adult COVID-19 patients included in the study, grouped by favipiravir administration.

| PARAMETER | Total<br>(n=150) | FVP cohort<br>(n=75) | Non-FVP cohort<br>(n=75) | p value |
| --- | --- | --- | --- | --- |
| Disease progression (n, %) | 27 (18.0) | 17 (22.7) | 10 (13.3) | 0.13 |
| 14-day all-cause mortality (n, %) | 19 (12.7) | 9 (12.0) | 10 (13.3) | 0.8 |
| Need for invasive mechanical ventilation (n, %) | 12 (8.0) | 8 (10.7) | 4 (5.3) | 0.22 |
| Need for immunomodulatory therapy (n, %) | 13 (8.7) | 10 (13.3) | 1 (1.3) | **<0.01** |
| Rate of PCR negativity (n, %) | 99 (66.0) | 55 (73.0) | 43 (57.3) | 0.05 |
| Time from diagnosis to disease progression<br>(days, median±IQR, min–max) | 7.0±7.0 (1.0–14.0) | 8.0±9.0 (3.0–14.0) | 4.5±9.8 (1.0–12.0) | 0.08 |
| Time from diagnosis to exitus<br>(days, median±IQR, min–max) | 12.0±7.5 (1.0–50.0) | 16.0±14.0 (6.0–50.0) | 8.5±10.3 (2.0–14.0) | **0.03** |
| Time from diagnosis to mechanical ventilation<br>(days, median±IQR, min–max) | 4.5±5.3 (1.0–12.0) | 5.5±5.3 (3.0–12.0) | 4.5±3.5 (1.0–12.0) | 0.79 |
| Time from diagnosis to immunomodulatory therapy<br>(days, median±IQR, min–max) | 7.0±5.5 (3.0–14.0) | 7.0±6.0 (3.0–14.0) | 4.0±0 (4.0–4.0) | 1.0 |
| Time from positive PCR to PCR negativity<br>(days, median±IQR, min–max) | 18.5±10.5 (3.0–55.0) | 22.0±15.5 (3.0–55.0) | 13.0±12.0 (3.0–32.0) | **<0.01** |
| LOS (days, median±IQR, min–max) | 15.0±10.8 (2.0–49.0) | 18.5±15.5 (3.0–49.0) | 13.0±8.5 (2.0–46.0) | **<0.01** |
| ICU LOS (days, median±IQR, min–max) | 5.0±2.0 (1.0–25.0) | 5.0±2.0 (2.0–12.0) | 5.0±5.0 (1.0–25.0) | 0.79 |
| Supportive therapies needed<sup>a</sup> (n, %): |  |  |  |  |
| - Nasal cannula | 79 (52.7) | 38 (50.7) | 41 (54.7) | 0.62 |
| - Venturi mask | 41 (27.3) | 18 (24.0) | 23 (30.7) | 0.35 |
| - Non-invasive ventilation | 4 (2.7) | 4 (5.3) | 0 | 0.12 |
| - Vasopressor therapy | 11 (7.3) | 8 (10.7) | 3 (4.0) | 0.11 |
| - Renal replacement therapy | 1 (0.7) | 1 (1.3) | 0 | 0.31 |
| - Parenteral nutrition | 1 (0.7) | 1 (1.3) | 0 | 0.31 |
| - Prone positioning | 3 (2.0) | 0 | 3 (4.0) | 0.24 |
| - Any | 89 (59.3) | 43 (57.3) | 46 (61.3) | 0.73 |
| Antiviral therapies given (n, %): |  |  |  |  |
| - Favipiravir | 75 (50.0) | 75 (100.0) | 0 | **<0.01** |
| - Chloroquin/hydroxychloroquin | 66 (44.0) | 5 (6.7) | 61 (81.3) | **<0.01** |
| - Oseltamivir | 34 (22.7) | 0 | 34 (45.3) | **<0.01** |
| - Lopinavir/ritonavir | 7 (4.7) | 0 | 7 (9.3) | **0.01** |
| - Remdesivir | 0 | 0 | 0 | n.a. |
| - Any<sup>b</sup> | 139 (92.7) | 75 (100.0) | 64 (85.3) | **<0.01** |
| Antibacterial therapies given (n, %): |  |  |  |  |
| - Ampicillin | 1 (0.7) | 1 (1.3) | 0 | 0.31 |
| - Amoxicillin/clavulanate | 5 (3.3) | 1 (1.3) | 4 (5.3) | 0.17 |
| - Cefazolin | 3 (2.0) | 3 (4.0) | 0 | 0.24 |
| - Ceftriaxon | 41 (27.3) | 13 (17.3) | 28 (37.3) | **<0.01** |
| - Cefepim | 2 (1.3) | 1 (1.3) | 1 (1.3) | n.a. |
| - Piperacillin/tazobactam | 20 (13.3) | 13 (17.3) | 7 (9.3) | 0.14 |
| - Meropenem | 9 (6.0) | 7 (9.3) | 2 (2.7) | 0.85 |
| - Imipenem/cilastatin | 15 (10.0) | 12 (16.0) | 3 (4.0) | 0.01 |
| - Vancomycin | 3 (2.0) | 3 (4.0) | 0 | 0.08 |
| - Linezolid | 2 (1.3) | 2 (2.7) | 0 | 0.15 |
| - Azithromycin | 63 (42.0) | 19 (25.3) | 44 (58.7) | **<0.01** |
| - Clarithromycin | 1 (0.7) | 1 (1.3) | 0 | 0.31 |
| - Levofloxacin | 13 (8.7) | 1 (1.3) | 12 (16.0) | **<0.01** |
| - Doxycyclin | 10 (6.7) | 0 | 10 (13.3) | **<0.01** |
| - Gentamicin | 4.0 (2.7) | 4 (5.3) | 0 | **0.04** |
| - Any<sup>b</sup> | 108 (72.0) | 47 (62.7) | 61 (81.3) | **0.01** |
| Immunomodulatory therapies given (n, %): |  |  |  |  |
| - Tocilizumab | 10 (6.7) | 9 (12.0) | 1 (1.3) | **0.01** |
| - Ruxolitinib | 0 | 0 | 0 | n.a. |
| - Baricitinib | 0 | 0 | 0 | n.a. |
| - Intravenous immunoglobulin | 2 (1.3) | 2 (2.7) | 0 | 0.49 |
| - Reconvalescent plasma | 0 | 0 | 0 | n.a. |
| - Systemic corticosteroid | 3 (2.0) | 3 (4.0) | 0 | 0.24 |

<sup>a</sup> Excluding invasive mechanical ventilation.
<sup>b</sup> Alone or in combination.
n.a. Not applicable.

**Table 3.** Independent predictors of disease progression among adult COVID-19 patients included in the study, grouped by progression occurence

|  | Disease progression<br>(n=27) | No disease progression<br>(n=123) | Univariate analysis |  | Multivariate analysis |  |
| --- | --- | --- | --- | --- | --- | --- |
|  |  |  | OR (95% CI) | p value | OR (95% CI) | p value |
| Age | 74.0±23.1 (41.0–91.0) | 66.0±12.9 (20.0–93.0) | 1.04 (1.01–10.9) | 0.01 | – |  |
| Male gender | 12 (44.0) | 64 (52.0) | 0.74 (0.32–1.72) | 0.47 |  |  |
| Time from disease onset to antiviral therapy | 3.3±7.8 (1–34) | 3.5±7.8 (1–34) | 0.91 (0.81–1.01) | 0.07* | n.a. |  |
| Essential hypertension | 19 (70.4) | 76 (61.8) | 1.64 (0.64–4.17) | 0.3 |  |  |
| Chronic heart disease | 18 (66.7) | 34 (27.6) | 5.81 (2.32–14.70) | <0.01 | 4.27 (1.41–12.98) | 0.01 |
| Chronic cerebral disease | 5 (18.5) | 19 (15.4) | 1.29 (0.43–3.84) | 0.64 |  |  |
| Diabetes mellitus | 10 (37.0) | 23 (18.7) | 2.69 (1.08–6.71) | 0.03 | – |  |
| Need for oxygen supportation | 16 (59.3) | 32 (26.0) | 4.13 (1.74–9.80) | 0.01 | – |  |
| Severe COVID-19 | 26 (96.3) | 50 (27.0) | 38.5 (5.0–333.30) | <0.01 | 21.28 (2.32–200.0) | <0.01 |
| Any supportive therapy needed | 26 (96.3) | 63 (51.2) | 5.10 (1.82–14.30) | <0.01 | – |  |
| Any antibacterial therapy needed | 22 (81.5) | 86 (69.9) | 1.89 (0.67–5.37) | 0.23 |  |  |
| Any antiviral therapy needed | 25 (92.6) | 114 (92.7) | 1.01 (0.21–4.98) | 0.98 |  |  |
| Treatment with favipiravir | 17 (63.0) | 58 (47.2) | 1.91 (0.81–4.48) | 0.14 | – |  |

\*The parameter was not included in the final model as co-linearity was not proven by the *Box–Tidwell* test with *Bonferroni's post hoc* correction ( $p < 0.01$ )

Although supportive therapies were needed in equivalent rates (43/75, 57.3% vs. 46/75, 61.3%, p=0.73), hospital LOS (18.5±16.0 vs. 13.0±9.0 days, <0.01) was higher with similar ICU admissions (12/75, 16.0% vs. 5/75, 6.7%, p=007). In the non-FVP cohort, patients mostly received chloroquin or hydroxychloroquin (5/75, 6.7% vs. 61/75, 81.3%, p<0.01). Preferred antibiotics were azithromycin (19/75, 25.3% vs. 44/75, 58.7%, p<0.01) and ceftriaxon (13/75, 17.3% vs. 28/75, 37.3%, p<0.01). Tocilizumab was given frequently to patients with cytokine storm (9/75, 12.0% vs. 1/75, 1.3%, p=0.01).

## 4. DISCUSSION

### 4.1 Present study

We performed a single-center, prospective observational, sequential cohort study by enrolling 150 hospitalized adult patients with moderate to severe COVID-19, receiving either favipiravir or other antiviral medications with standard of care during the first 4 months of SARS-CoV-2 pandemic in Hungary. There was no statistically significant differences in time to antiviral therapy initiation from PCR positivity, COVID-19 disease severity, need for oxygen supportation and ICU admittance rates between cohorts. Disease progression, 14-day all-cause mortality, need for invasive mechanical ventilation and PCR negativity rate seemed to be unaffected by favipiravir exposure compared to other antivirals, and there was higher demand for immunomodulatory therapy administration among patients in the favipiravir cohort.

### 4.2 Previous studies

Pharmacological approaches, as well as clinical studies describing favipiravir treatment strategies for COVID-19 patients were reported in the literature. Although favipiravir demonstrated good *in vitro* inhibitory activity against SARS-CoV-2, the optimal pharmacological dose for COVID-19 treatment has yet to be determined, as the dose recommendations (as used in our study) based on pharmacokinetic experiments and clinical trials. Data of studies involving patients with other severe viral infections (influenzavirus, Ebolavirus) might be insufficient to maintain an adequate serum concentration, especially in critically ill patients [2, 11, 12]. Although a review found that favipiravir has a favourable safety profile concerning serious adverse events, the main side effects are hyperuricaemia, teratogenicity and QTc prolongation. Establishment of long-term safety profile among COVID-19 patients needs more pharmacovigilance data [13].

According to data from the literature, clinical usefulness of favipiravir in COVID-19 may somewhat be limited. A prospective clinical trial randomizing 240 adult patients in an 1:1 fashion with clinically confirmed COVID-19 to conventional therapy and umifenovir or favipiravir reported that although favipiravir administration associated with shorter time to defervescence and diminishment of cough, the drug could not significantly improve 7-day clinical recovery rate as a primary endpoint. Furthermore, rates of noninvasive mechanical ventilation, supplementary oxygen demand or all-cause mortality did not show differences between groups [14]. An open-labeled non-randomized study conducted by matching 35 microbiologically confirmed COVID-19 patients treated with favipiravir and 45 patients receiving lopinavir/ritonavir (all with additional interferon-alpha inhalation), within one week after onset of symptomps. Authors documented statistically higher improvement rates in chest imaging and faster viral clearance among patients receiving favipiravir, but effect of favipiravir on disease progression or mortality rates were not reported [15]. A recently published multicenter phase II/III randomized trial enrolled patients with moderate COVID-19 within a median of 6.7 days from start of symptomps. The primary efficacy endpoint was the elimination of SARS-CoV-2 by Day 10. On day 5, viral clearance was significantly more prevalant in the favipiravir arm (p=0.018), but on day 10 the difference was statistically non-significant (p=0.155). Authors conclude that in their pilot study, favipiravir appeared beneficious among moderately ill patients [16]. In small case series studies, favipiravir was administered with nafamostat mesylate or methylprednisolone for COVID-19 patients in different disease stages, but due to the lack of control groups and antiviral combination usage, the extent favipiravir effect on clinical cure remains ambiguous [17-19]. In all, we think that our findings could be reflected by data from current literature, and the role for favipiravir should not be overestimated in the treatment of adults with moderate to severe COVID-19.

### 4.3 Limitations

Our study had several limitations. As the pandemic is still ongoing, knowledge about COVID-19 is changing fast, and despite best efforts, *per protocol* treatment allocation might have been lagging behind literature evidence. National drug availability might have biased treatment allocation. The initiation of alternative antiviral medications might have been biased by (contra)indication. Age and comorbidity differences between our cohorts could represent the temporal progression of the epidemic in Hungary: younger people were affected sooner in larger numbers, and this might have confounded some baseline characteristics due to the sequential cohort design. The number of included patients is relatively low, however, an exact *a priori* study size calculation was not feasible due to study design and enrollment. Chest CT was not readily obtainable from some patients due to technical reasons.

## 5. CONCLUSION

Among adult patients hospitalized with COVID-19, an overall beneficial effect of favipiravir on disease progression could not be proven in this study. Due to limitations, further prospective trial data are needed to elucidate the role of favipiravir in COVID-19 treatment.

## Data Availability

Anonymised data of patients are available from the corresponding author on reasonable request.

## Acknowledgements

The authors would like to thank the healthcare workers of our centre for their sacrifice during these times. Together, we are stronger. Preliminary results of the study were presented at ECCVID 2020 (September 23–25, 2020).

## Access to data

Anonymised data of patients are available from the corresponding author on reasonable request.

## Authors’ contributions

BGSZ and LKSZ contributed equally to the manuscript (*in equo loco*). BGSZ: management of patients, data collection, data analysis, preparation of study protocol, preparation of the manuscript; LKSZ: management of patients, data collection, data analysis, preparation of study protocol, preparation of the manuscript; BP: data collection, data analysis; ZSG: data collection, data analysis; ZSB: management of patients, review of the manuscript; ZSB: management of patients, review of the manuscript; EB: management of patients, review of the manuscript; JB: management of patients, review of the manuscript; EC: management of patients, review of the manuscript; KF: management of patients, review of the manuscript; AH: management of patients, review of the manuscript; NKD: management of patients, review of the manuscript; CSL: management of patients, review of the manuscript; KN: management of patients, review of the manuscript; JK: management of patients, review of the manuscript; ELN: management of patients, review of the manuscript; AO: management of patients, review of the manuscript; EP: management of patients, review of the manuscript; AR: management of patients, review of the manuscript; JSZ: management of patients, review of the manuscript; BSZ: management of patients, review of the manuscript; ASZ: management of patients, review of the manuscript; SZT: management of patients, review of the manuscript; ZSV: management of patients, review of the manuscript; OW: management of patients, review of the manuscript; JSZ: management of patients, review of the manuscript, COVID-19 Team Leader; BL: management of patients, preparation of study protocol, preparation and review of the manuscript.

## Ethics approval

The Institutional Review Board of South Pest Central Hospital, National Institute of Hematology and Infectious Diseases (Budapest, Hungary) approved the study protocol. Approval for the use of off-label investigational drugs was granted by our institutional review board, as well as the National Institute of Pharmacy and Nutrition in relation to the COVID-19 surge in Hungary *(ogyei*.*gov*.*hu/tajekoztato_a_veszelyhelyzet_megszunesevel_kapcsolatos_a_covid_19_jarvany _idejen_kulonos_meltanylast_erdemlo_betegellatasi_erdekhez_kotheto_gyogyszeralkalmazas ok_bejelenteserol)*.

## ABBREVIATIONS

ARDS: acute respiratory distress syndrome
BAL: bronchoalveolar lavage
COVID-19: coronavirus disease-19
CRP: C-reactive protein
CS: cytokine storm
CT: computed tomography
ECDC: European Centre for Disease Prevention and Control
EUCAST: European Committee on Antimicrobial Susceptibility Testing
FVP: favipiravir
ICU: intensive care unit
IQR: interquartile region
LDH: lactate dehydrogenase
LOS: length of stay
PCR: polymerase chain reaction
RCT: randomized clinical trials
SARS-CoV-2: severe acute respiratory syndrome coronavirus-2
SOC: standard of care
STROBE: Strengthening the Reporting of Observational studies in Epidemiology

## REFERENCES

[1] National Institutes of Health. COVID-19 Treatment Guidelines. 2020. https://www.covid19treatmentguidelines.nih.gov/whats-new/. (Accessed 2020.09.01).

[2] Shannon A, Selisko B, Le N, Huchting J, Touret F, Piorkowski G, et al. Favipiravir strikes the SARS-CoV-2 at its Achilles heel, the RNA polymerase. bioRxiv 2020.

[3] Coomes EA, Haghbayan H. Favipiravir, an antiviral for COVID-19? J Antimicrob Chemother 2020;75:2013–4.

[4] European Centre for Disease Prevention and Control. Case definition for coronavirus disease 2019 (COVID-19), as of 29 May 2020. 2020. https://www.ecdc.europa.eu/en/covid-19/surveillance/case-definition. (Accessed 2020.09.01).

[5] World Health Organization. Clinical management of COVID-19. 2020. https://www.who.int/publications/i/item/clinical-management-of-covid-19. (Accessed 2020.09.01.).

[6] Force ADT, Ranieri VM, Rubenfeld GD, Thompson BT, Ferguson ND, Caldwell E, et al. Acute respiratory distress syndrome: the Berlin Definition. JAMA 2012;307:2526–33.

[7] Mehta P, McAuley DF, Brown M, Sanchez E, Tattersall RS, Manson JJ. COVID-19: consider cytokine storm syndromes and immunosuppression. Lancet 2020;395:1033–4.

[8] Emberi Erőforrások Minisztériuma. A 2020. évben azonosított új koronavírus (SARS-CoV-2) okozta fertőzések (COVID-19) megelőzésének és terápiájának kézikönyve. 2020. https://koronavirus.gov.hu/sites/default/files/sites/default/files/imce/magyar_koronavirus_kezikonyv_2020.06.25.pdf . (Accessed 2020.09.01.) [In Hungarian].

[9] Metlay JP, Waterer GW, Long AC, Anzueto A, Brozek J, Crothers K, et al. Diagnosis and Treatment of Adults with Community-acquired Pneumonia. An Official Clinical Practice Guideline of the American Thoracic Society and Infectious Diseases Society of America. Am J Respir Crit Care Med 2019;200:e45–e67.

[10] von Elm E, Altman DG, Egger M, Pocock SJ, Gøtzsche PC, Vandenbroucke JP. The Strengthening the Reporting of Observational Studies in Epidemiology (STROBE) statement: guidelines for reporting observational studies. Lancet 2007;370:1453–7.

[11] Irie K, Nakagawa A, Fujita H, Tamura R, Eto M, Ikesue H, et al. Pharmacokinetics of Favipiravir in Critically Ill Patients With COVID-19. Clin Transl Sci 2020;13:880–5.

[12] Du YX, Chen XP. Favipiravir: Pharmacokinetics and Concerns About Clinical Trials for 2019-nCoV Infection. Clin Pharmacol Ther 2020;108:242–7.

[13] Pilkington V, Pepperrell T, Hill A. A review of the safety of favipiravir – a potential treatment in the COVID-19 pandemic? J Virus Erad 2020;6:45–51.

[14] Chen C, Zhang Y, Huang J, Yin P, Cheng Z, Wu J, et al. Favipiravir versus Arbidol for COVID-19: A Randomized Clinical Trial. medRxiv 2020.

[15] Cai Q, Yang M, Liu D, Chen J, Shu D, Xia J, et al. Experimental Treatment with Favipiravir for COVID-19: An Open-Label Control Study. Engineering (Beijing) 2020.

[16] Ivashchenko AA, Dmitriev KA, Vostokova NV, Azarova VN, Blinow AA, Egorova AN, et al. AVIFAVIR for Treatment of Patients with Moderate COVID-19: Interim Results of a Phase II/III Multicenter Randomized Clinical Trial. Clin Infect Dis 2020.

[17] Murohashi K, Hagiwara E, Kitayama T, Yamaya T, Higa K, Sato Y, et al. Outcome of early-stage combination treatment with favipiravir and methylprednisolone for severe COVID-19 pneumonia: A report of 11 cases. Respir Investig 2020.

[18] Doi K, Ikeda M, Hayase N, Moriya K, Morimura N, Group C-US. Nafamostat mesylate treatment in combination with favipiravir for patients critically ill with Covid-19: a case series. Crit Care 2020;24:392.

[19] Yamamura H, Matsuura H, Nakagawa J, Fukuoka H, Domi H, Chujoh S. Effect of favipiravir and an anti-inflammatory strategy for COVID-19. Crit Care 2020;24:413.

